# Unraveling the COVID-19 hospitalization dynamics in Spain using publicly available data

**DOI:** 10.1101/2021.09.03.21263086

**Authors:** Alberto Aleta, Juan Luis Blas-Laína, Gabriel Tirado Anglés, Yamir Moreno

## Abstract

**Background:** One of the main challenges of the ongoing COVID-19 pandemic is to be able to make sense of available, but often heterogeneous and noisy data, to characterize the evolution of the SARS-CoV-2 infection dynamics, with the additional goal of having better preparedness and planning of healthcare services. This contribution presents a data-driven methodology that allows exploring the hospitalization dynamics of COVID-19, exemplified with a study of 17 autonomous regions in Spain.

**Methods:** We use data on new daily cases and hospitalizations reported by the Ministry of Health of Spain to implement a Bayesian inference method that allows making short and mid-term predictions of bed occupancy of COVID-19 patients in each of the autonomous regions of the country.

**Findings:** We show how to use given and generated temporal series for the number of daily admissions and discharges from hospital to reproduce the hospitalization dynamics of COVID-19 patients. For the case-study of the region of Aragon, we estimate that the probability of being admitted to hospital care upon infection is 0·090 [0·086-0·094], (95% C.I.), with the distribution governing hospital admission yielding a median interval of 3·5 days and an IQR of 7 days. Likewise, the distribution on the length of stay produces estimates of 12 days for the median and 10 days for the IQR. A comparison between model parameters for the regions analyzed allows to detect differences and changes in policies of the health authorities.

**Interpretation:** The amount of data that is currently available is limited, and sometimes unreliable, hindering our understanding of many aspects of this pandemic. We have observed important regional differences, signaling that to properly compare very different populations, it is paramount to acknowledge all the diversity in terms of culture, socio-economic status and resource availability. To better understand the impact of this pandemic, much more data, disaggregated and properly annotated, should be made available.

## Introduction

During the early stages of the COVID-19 pandemic, one of the greatest public health concerns was the adequacy of healthcare resources to treat infected cases. With cases growing exponentially, and roughly 10% of the detected cases needing hospitalization in intensive care units, it was soon realized that national health systems could be easily overwhelmed.^1–3^. To slow the advance of the epidemic and protect national health systems, many countries implemented strict lockdowns during the first wave of the epidemic. Further waves also forced the application of important non-pharmaceutical interventions, reducing the effective reproduction rate of the disease and preventing the collapse of the healthcare systems.^4–7^

At the same time, this event is the largest pandemic in the Digital Age, with wide access to the internet and ubiquity of social networks, representing a completely paradigm shift in terms of communication, data collection, storage and dissemination at all scales.^8,9^ Yet, many healthcare systems were ill-prepared for an event of such scale. For instance, a group of researchers, journalists and non-profit organizations studied the COVID-19 data dashboards for each state in the US and concluded that the US lacked “standards for state-, county-, and city-level public reporting of this life-and-death information”, calling it an “information catastrophe”.^10^ Many numbers had important lags, with a quarter of deaths reported less than six days after they had occurred while another 25 percent were reported more than 45 days later.^11^ Furthermore, even if open data was available, it often lacked the granularity necessary to understand the gender, racial, ethnic and economic disparities created or amplified by the pandemic.^12^

Similar problems can also be found in many countries in the EU.^13^ In the case of Spain, during the first wave, the reporting delay was highly variable, ranging from a few days in early March to over two weeks in April.^14^ The problem was further exacerbated due to the high decentralization of the health system in Spain, managed by each autonomous region independently, which caused many problems of data synchronization and case definition.^15^ In March 2021, already one year into the pandemic, a report on child mortality of COVID-19 in Europe showed that Spain had the highest mortality of all the countries analyzed.^16^ A deeper investigation revealed that some patients over 100 years old had been incorrectly labeled as underaged, resulting in an abnormal number of children deaths which was later amended, yielding results compatible with other European countries.^17^

Thus, it is important to devise methodologies that can fully exploit this public data sources and detect errors, inconsistencies or underlying methodological changes that are not correctly addressed by the providers. In this paper, we focus on the context of hospital dynamics and present a simple model to translate detected incidence into hospital admissions and bed occupancy. The model was first implemented to estimate bed occupancy in the autonomous region of Aragon during the fourth wave of the pandemic, but it is here applied to study the temporal dynamics of hospitalizations of other regions of the country too. Besides, we also show that this tool can also be used to detect changes underlying the data which cannot be easily detected otherwise. Lastly, we apply the methodology to compare the evolution of the 17 autonomous regions in Spain and relate it to the testing policy implemented in each region. Our methodology can therefore be used to scrutinize the unfolding of the disease and its impact, together with the NPIs adopted, on hospital occupancy. As a result, it can also be employed as a tool for preparedness and healthcare planning, alleviating the high and yet-to-assess delays in diagnosis and treatment of other -potentially deadly, e.g., cancer, tuberculosis, etc-diseases caused by the pandemic.

## Methods

### Overview

The model estimates daily admissions and bed occupancy based on the daily incidence data reported by the authorities. To do so, we estimate the probability of being admitted into a hospital upon being confirmed as a positive case, as well as the delay between detection and admission, and the length of stay (LoS). We employ a Bayesian inference model trained on data from July 2020 to late November 2020. With this information, we can reproduce the hospital dynamics in the autonomous region of Aragon - taken here as a case study-from December 2020 to June 2021, and produce forecasts on hospitalization based on any technique for predicting the daily incidence. In addition, we show the generality of the methodology by applying it to all the autonomous regions of Spain. This allows us to study their evolution, and analyze their differences, strategies for case detection and the effects of the first stages of the vaccination campaigns.

### Datasets

We use the open data provided by the Spanish Health Ministry, which reports the number of confirmed cases by symptom onset, or notification date minus 3 days if it is not available, and the number of daily hospital admissions.^18^ We further complement this dataset with the one provided by the regions of Aragon and Catalonia for some specific analysis.^19,20^ In particular, the main dataset provided by the Spanish Health Ministry does not contain information on discharges or bed occupancy, limiting regional comparisons to new admissions.

There is an alternative repository focused on hospitalization but has several gaps since it is not updated on Saturdays, Sundays and special holidays.^21^ Furthermore, the data on discharges “might not be collected exhaustively (sic)”. Thus, for the analysis involving bed occupancy, we resort to the regional repository. Note also that the quality, accessibility and depth of each regional repository varies greatly, and not always matches the data provided by the Ministry, as we show later for the case of Catalonia. Another example is the incidence data provided by the government of Aragon, which is based on the notification date, while the incidence data provided by the Spanish Health Ministry is based on symptoms’ onset. Therefore, comparisons among different regions are performed using the standardized data provided by the Spanish Health Ministry.

### Model

For each region *r*, given the number of daily cases on day *i*, 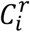, we estimate the daily number of hospital admissions at time *t* as

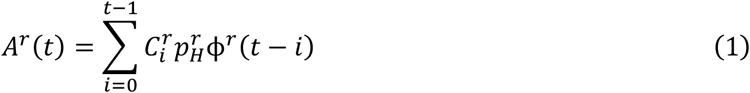

where 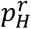 is the regional probability of hospitalization and *ϕ*^*r*^(*t* − *i*) is a delay function that yields the probability of being admitted to the hospital on day *t* given that the case was notified on day *i*. Data on the characteristics of all hospitalized individuals between early August and late November provided by the Health Department of the region of Aragon indicates that *ϕ* might be approximated by a Half-Cauchy distribution:

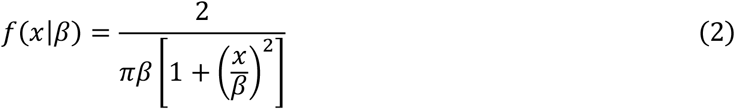

Similarly, to determine the daily number of discharges, we can apply a convolution between the number of daily admissions and the distribution of the length of stay, ψ,

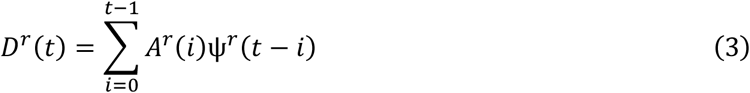

In this case, the data showed that ψ might be well approximated by a log-normal distribution:

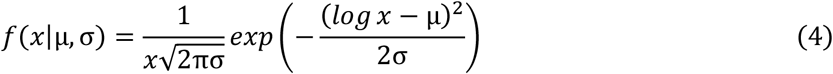

So that the bed occupancy in region *r*, on day *t*, given the daily incidence up to that day will be given by

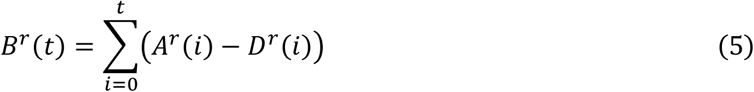

### Bayesian inference

To estimate the parameters, we implement a Bayesian statistical model using PyMC3.^22^ Note that all parameters cannot be estimated simply from bed occupancy data (eq. (5)) since a larger probability of hospitalization with small LoS can yield the same occupancy as a small probability of hospitalization with large LoS. Thus, we first estimate the set of model parameters for the daily admissions, 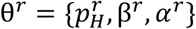, using Bayesian inference with Markov-chain-Monte-Carlo (MCMC). We assume that the likelihood of observing the real-world data point, *Â*^*r*^(*t*), is given by NegBinomial(μ = *A*^*r*^ (*t*|θ), α) to allow for over-dispersion. We run 4 independent chains using NUTS with 2000 burn-in steps and 5000 steps to approximate the posterior distribution of the parameters with uninformed priors.

With these parameters and eq. (1) we can obtain the daily number of admissions as a function of the number of new detected cases. This information, together with eq. (5) and the observed bed occupancy, 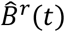, can be used to estimate the parameters of the log-normal distribution governing discharges. In this case we assume that the likelihood of observing the real data point is given by Normal(μ = *B*^*r*^(*t*|θ), σ), but otherwise the training procedure is the same as for the admissions.

### Forecasting

The previous model can translate new infections into new admissions, use the distribution of LoS and, finally, estimate bed occupancy for the short-term (2-3 days). Thus, to increase the prediction window, it is necessary to forecast future incidence. There are many techniques available to do this, ranging from classical compartmental models to Bayesian inference akin to the one used here, detailed agent-based models, metapopulation models, branching processes, machine learning and deep learning techniques or other time-series forecasting tools.^23–29^ Since this is not the main purpose of this work, we resort to a simple heuristic that yields satisfactory results and is easier to communicate to non-technical stakeholders.

First, we computed the evolution of the effective reproduction number in the region of Aragon from early July to mid-November. Then, we explored how this quantity changed from day to day by dividing the value at time *t, R*(*t*), over its value on the previous day, *R*(*t* − 1). This analysis revealed that during the growing phase of an outbreak, the daily increase on *R*(*t*) is below 5%. Similarly, during the shrinking phase induced by the non-pharmaceutical interventions imposed by the authorities, the daily decrease is close to 5%, see figure S3. Given this observation, we can forecast the value of the reproduction number as

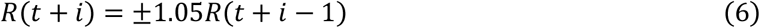

with the sign chosen depending on whether the outbreak is growing or decreasing. Then, we can estimate the number of new cases by multiplying this quantity iteratively by the number of cases observed or estimated in the previous days, and introduce it in eq. (5) to forecast bed occupancy.

## Results

### Estimating model parameters for daily admissions

To estimate the model parameters, we choose the period between July 1, right before the second wave of infections started in Spain, and December 1, right before the beginning of the Christmas wave. In figure 1, we show the number of daily admissions obtained using equation (1), while in figure S1 we depict the corresponding prior and posterior distributions. The period before December 1 can be regarded as the training period for the model. Afterwards, the prediction is labeled “informed forecast” since it requires knowledge of the number of new cases.

**Figure 1.**
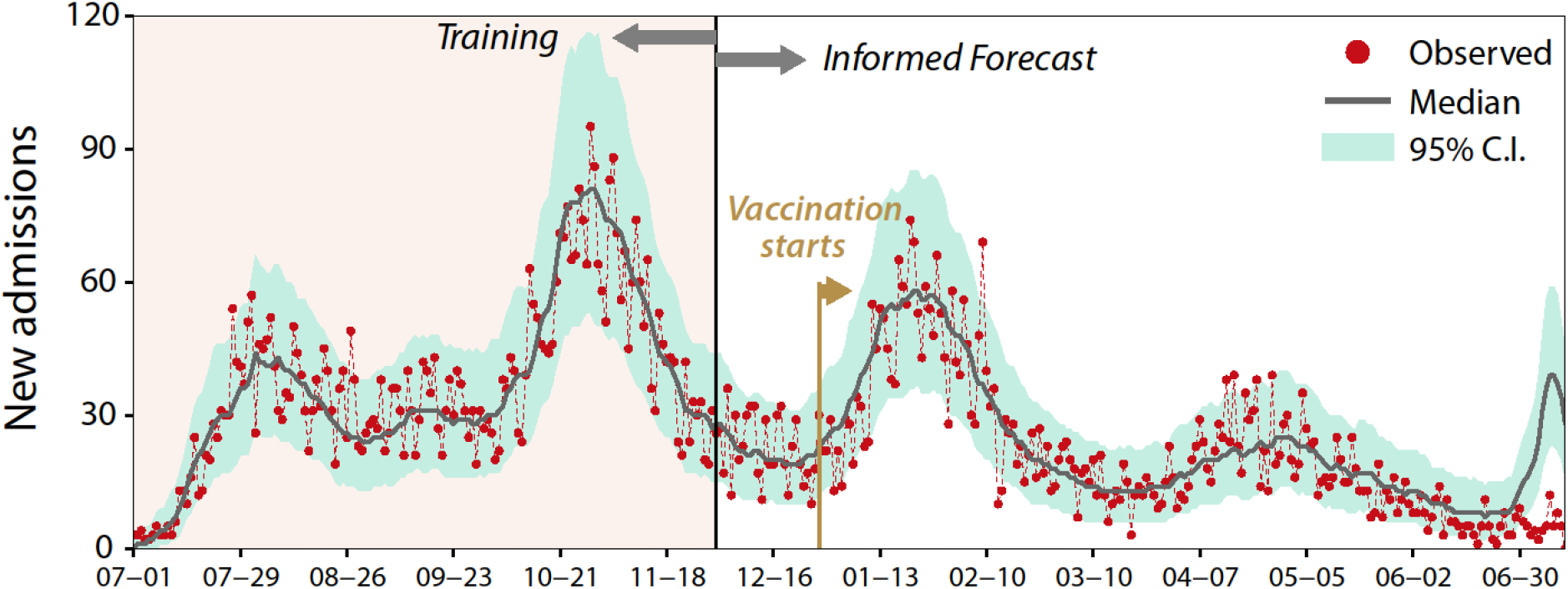
Number of daily admissions to hospitals in the region of Aragon. To estimate the parameters of the model, we use the information available up to December 1, 2020. From December 1 the number of admissions is obtained using the estimated parameters and the observed number of daily detected cases, applying eq. (1). Note that on December 28 the vaccine roll-out started in Spain. The estimated parameters are 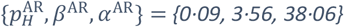.

A few remarks are in order. First, the region of Aragon was hardly hit during the training period, with two infection waves, while other Spanish regions only had one. Second, the number of daily admissions can vary substantially, yielding a large prediction credible interval. Third, we observe that the prediction is unaffected by vaccination until late May since the probability of hospitalization estimated in summer 2020 still gives satisfactory results until that date. Lastly, there is a new wave of infections in early summer 2021 that has been associated with individuals between 15 and 25 y.o. because of several parties hosted to celebrate the end of the academic year. This important difference in the age profile explains why the incidence is not translated into hospital admissions. Note also that this group, up to those dates, was excluded from (i.e., not prioritized) vaccination.

There can be several explanations for the small impact vaccination had on hospitalization dynamics during the first half of the year. First, early vaccination was mainly focused on the most vulnerable, who might not have had the chance to go to the hospital if they developed severe infection. Thus, reducing infections among that small group had a minor impact in both cases and hospitalizations. Second, another possible explanation is that vaccination reduced the severity of cases, hindering case detection. Indeed, since vaccination reduces the chance of suffering severe disease, it is possible that those infections might have been undetected, not contributing to lower the value of *p*_*H*_. Lastly, it has been proposed that the profile of hospitalized cases shifted towards un-vaccinated groups, compensating the effect of vaccination and leaving *p*_*H*_ unaltered. Surveillance data shows that, indeed, the fraction of hospitalizations corresponding to the oldest age groups (older than 80 y.o.) has steadily declined since vaccination started. Conversely, the hospitalization rate within the 40 to 49 and 50 to 59 y.o. groups increased (see Fig S4), especially in May. Note that the vaccination of these groups started precisely in May, which explains the observed pattern. Recently, daily incidence data disaggregated by age has been made publicly available by the Ministry. Further studies should use this data to better estimate the hospitalization dynamics by age-group and its relationship with vaccination.

### Estimating model parameters for daily discharges

In figure 2, we show the observed daily occupancy of hospital beds in Aragon together with the estimated one. We observe that the agreement is very good, even after vaccination started, until the beginning of April 2021. From that point on, the estimated occupancy is lower than the observed one. Given that the estimated admissions still agree with the observed values of this variable, this implies that the length of stay for the age groups admitted to the hospital during those dates is larger than the average. Whether this is the effect of protecting the oldest individuals from severe infection, or due to the circulation of new variants, is something that cannot be addressed unless more disaggregated data is released (e.g., discharges by age-group).

**Figure 2:**
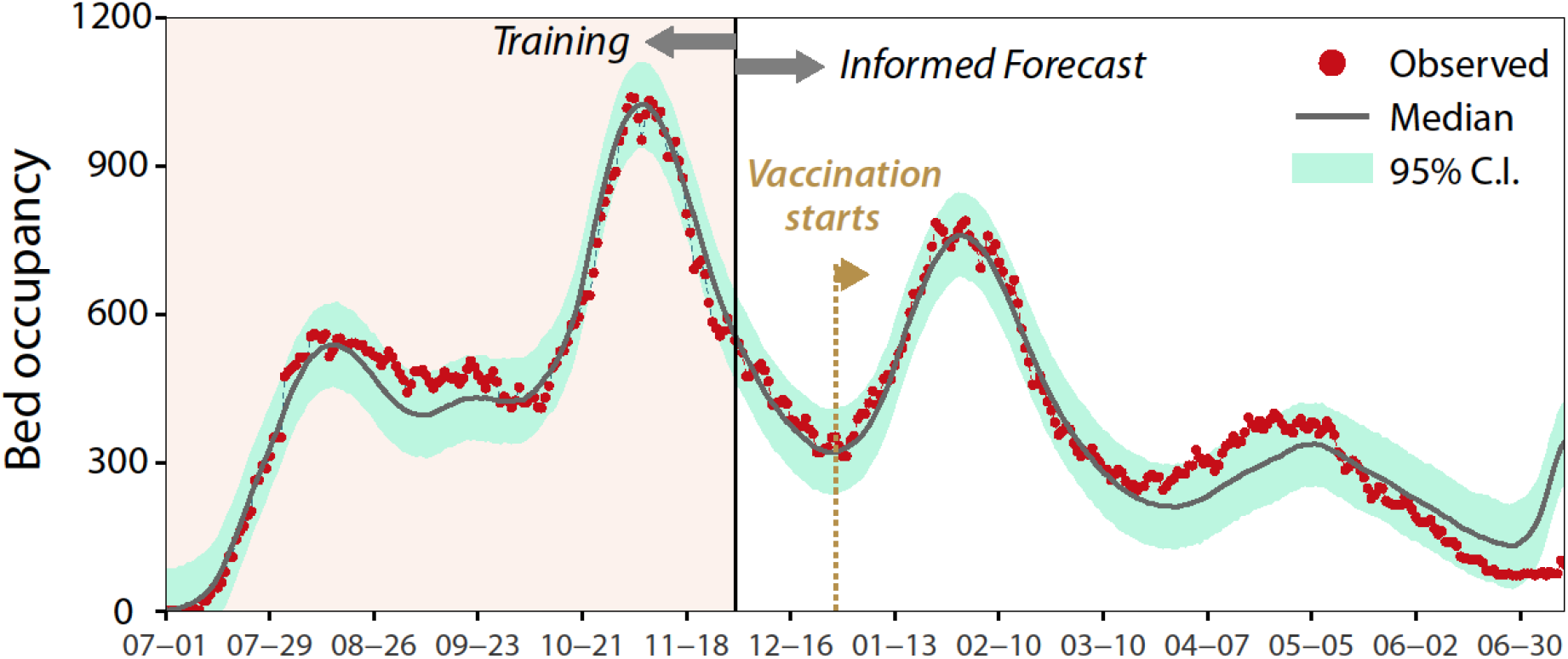
Daily number of beds occupied by COVID-19 patients in Aragon. To estimate the parameters of the model, we used information up to December 1, 2020. From December 1, the occupancy is obtained using the number of new detected cases, together with equations (1) and (5). Note that on December 28 vaccine roll-out started. The estimated parameters are 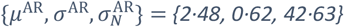.

### Bed occupancy in Aragon

In figure 3, we show the forecasts performed during the Christmas wave. Each Monday, the algorithm ran on the data collected up to that date and forecasted bed occupancy as a function of the expected change of *R*(*t*). Three scenarios were considered: pessimistic (+5%), neutral (+0%) and optimistic (−5%), which could be chosen depending on domain expertise and the unfolding of events, such as the imposition of new containment measures. A summary of all the estimated parameters used in the forecast is presented in Table S1.

**Figure 3:**
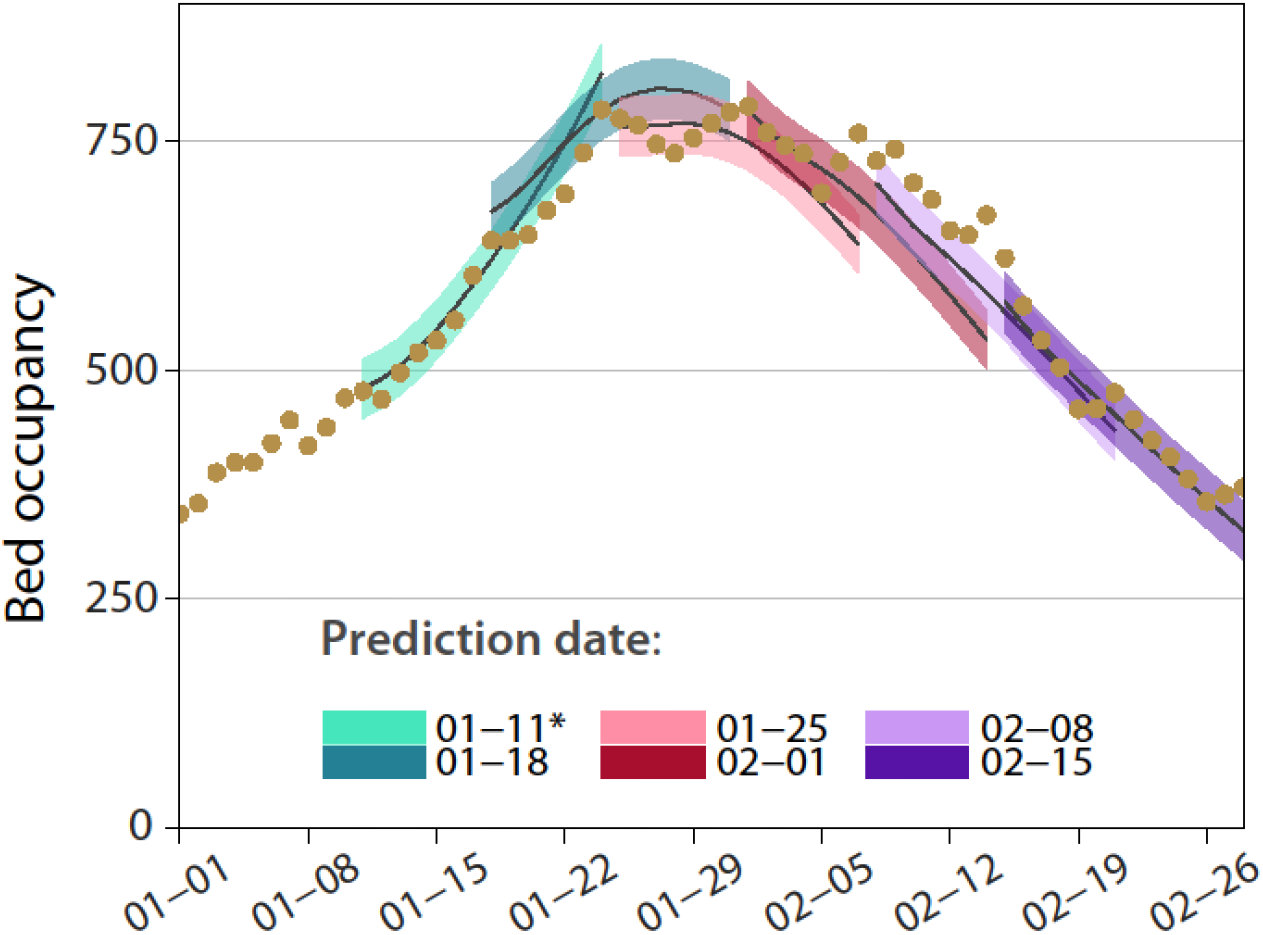
Forecasting bed occupancy in Aragon during the 2020-2021 Christmas wave. Dots show the actual value of bed occupancy, while shaded regions display the 95% CI of the forecast for the week starting at the indicated date. At each date depicted in the figure, the prediction algorithm runs on the number of cases detected up to that date and forecasts occupancy assuming a certain change of R(t). In the prediction made on January 11, 2021, the change is assumed to be +0%. Following the introduction of several containment measures on January 15, the daily change was assumed to be -5%.

We observe that, albeit the prediction of new cases is quite simplistic, the forecast of occupancy is remarkably good, especially during the first days of the prediction. In particular, the procedure can correctly estimate the maximum bed occupancy and the date when the tendency shifts downwards. Note also that this model can be applied to the output of any incidence prediction algorithm, and thus it can be easily adapted to other regions or more complex scenarios.

### Regional comparison of new admissions

The model can be used to compare the characteristics and temporal evolution of the outbreaks in each autonomous region of Spain. This comparison is especially interesting since each region manages its own healthcare service and response to the pandemic, which might lead to different values for the parameters of the model. Thus, we have implemented the model to estimate new admissions for each of the 17 autonomous regions in Spain (the 2 autonomous cities, Ceuta and Melilla, have been discarded due to their small size). In figure S5, we show the number of new daily admissions up to May 1, 2021, in each region. As in the previous case, we compute their corresponding parameters 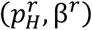 using data from July 1, 2020, to December 1, 2020. The model can estimate very well the number of new admissions in all regions, although there are some territories in which the estimation is slightly worse (see table S2).

A lower quality of the fit usually signals that the region changed its testing capacities, rather than other external factors such as emergence of new variants. For instance, in the Principality of Asturias, we observe that after mid-November the model tends to overestimate new admissions. The reason for this discrepancy can be that either some interventions reduced the value of *p*_*H*_, or that the surveillance system was improved, and more cases were detected. It turns out that Asturias was one of the three regions, together with the Canary Islands and the Balearic Islands, that did not use antigen tests in their surveillance system. This situation changed on November 10, 2020, which would explain why our predictions overestimate occupancy after mid-November 2020. A similar situation can be observed from mid-January 2021 in the Balearic Islands, which can probably be related to a massive testing campaign using antigen tests that started on mid-January 2021 and extended up to February 2021. Lastly, we also observe a slight overestimation of occupation in the region of Madrid. In this region, the health department of the autonomous community redefined contact tracing on September 28, 2020, excluding social encounters from the surveillance system. This change was reverted on November 20, 2020. The first change would force *p*_*H*_ to increase, while the latter would produce a larger number of detected cases. The combination of both would result in a larger estimation of bed occupancy from late November, therefore nicely explaining the divergence obtained in the model.

In figure 4, we present a summary of the 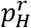 and β^*r*^ values obtained in the estimation of new admissions for all the autonomous regions of Spain. We observe that most regions cluster together around the same values, with a value for the probability of an infected individual needing hospital care upon detection around 9% and a delay of 3 to 4 days between detection and admission. The regions with most different values are those for which the prediction is worst, as previously discussed. It is interesting to mention that Catalonia (CT) has the lowest probability, which could indicate that the region is detecting more cases than the rest. However, as shown in figure S6, this is not indeed the case as the observed difference is a consequence of the data alone. Certainly, there are important differences between the values reported by the Spanish Health Ministry and the regional government of Catalonia. Using the values provided by the latter, the hospitalization probability is 8.5%, in line with the rest of the regions.

**Figure 4:**
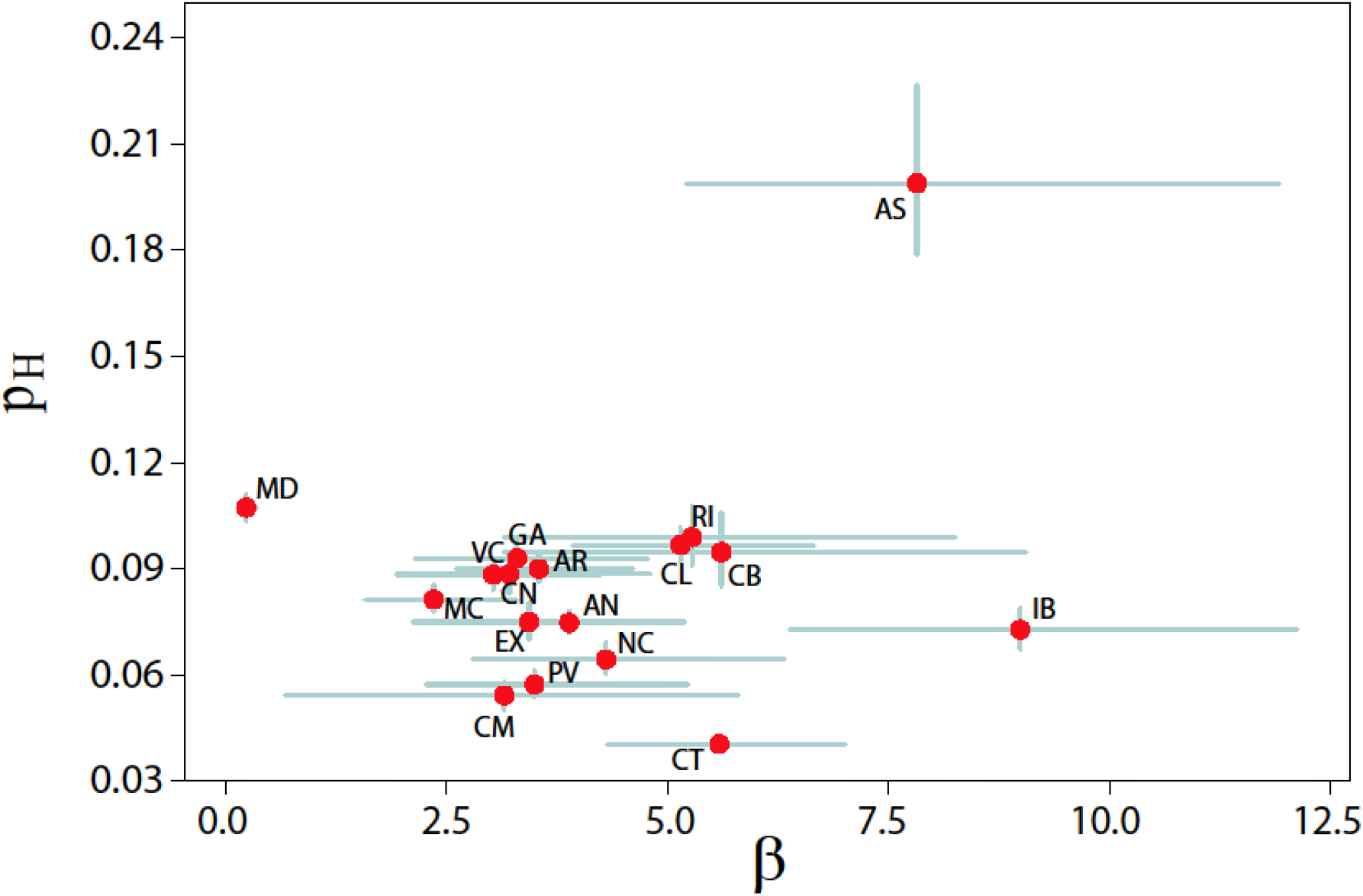
Admission dynamics in each region of Spain. Estimated value of the probability of being admitted into the hospital upon detection, p_H_, versus the parameters of the Half-Cauchy distribution governing the delay between detection and admission, β. The horizontal and vertical errorbars indicate the 95% C.I. Labels represent the ISO abbreviation of each autonomous region in Spain. The complete list of equivalences in shown in table S2.

Finally, it would also be possible to hypothesize that the observed differences among the regions in which the prediction was successful to the characteristics of their populations. For instance, one of the drivers of these differences could be that some regions have a larger fraction of the population within the eldest age-groups, which are more prone to need hospital care. However, in figure S7, we show that the correlation between these variables is very weak and cannot explain the difference.

## Discussion

COVID-19, the largest pandemic of the Digital Age, has revolutionized the way in which public health information is created, stored and shared. At the same time, it has highlighted the weaknesses of the information systems of many governmental and healthcare institutions. Even when the data is openly shared, the lack of information on how it was obtained, continuous changes in government policies, and availability of resources, makes it challenging to use the data and perform proper comparisons between different regions. This can be easily seen when data from different sources, or from the same source but for different observables, are compared.

The model presented here focuses on incidence and bed occupancy data. In particular, it reproduces the dynamics associated with hospitalization as a function of the number of detected cases. In the case of Aragon, we have obtained an admission probability of 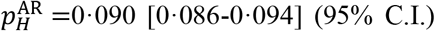 and the distribution governing hospital admission follows a Half-Cauchy distribution with β^AR^ =3·557[2·579-4·564] (95% C.I.). This distribution yields a median interval of 3·5 days and an IQR of 7 days, which is remarkably close to the estimated values based on individual data provided by the regional government (median 4 days, IQR 6 days). Note that small differences are expected since the way in which hospitals record case detection and admission might not be the same as the one provided by the Ministry of Health. For instance, while the Ministry of Health reports new cases as a function of symptoms’ onset, the regional government provides new cases based on notification date.

Similarly, the distribution on the length of stay can be approximated by a Log-Normal distribution with μ^AR^ =2·476 [2·441-2·512] (95% C.I.) and σ^AR^ =0·620 [0·554-0·688] (see figure S2 and table S1 for a summary of the estimated parameters). This distribution yields a median of 12 days with an IQR of 10 days. Data provided by the regional government shows that the median value is between 8 and 9 days with an IQR of 8 days for surviving patients, while for deceased patients the median LoS is 9 days and the IQR is 11 days. Note that in our model we do not distinguish the reason for discharge. Besides, even though this quantity is less prone to biases due to the quality of the surveillance system, some noise might be introduced depending on the discharge policy (for instance, it is uncommon to discharge patients during weekends). Nevertheless, these values are compatible with those provided by the regional authorities, as well as the ones found in the literature for other parts of the world.^30^

Importantly, the analysis shows that the situation is far from being static. During the course of the epidemic in Aragon, we have identified nontrivial changes in the LoS, and noticeable modifications on the detection capabilities in other regions of Spain. Furthermore, we have seen that there are important differences in the hospital dynamics of each region, and the quality and quantity of data provided by their institutions. Data for the same region, extracted from two different sources, can yield very different values, which might cast doubts on its reliability. Lastly, the lack of disaggregated data on very basic characteristics, such as age, limits the conclusions that can be obtained, such as the possible role of emerging variants. This problem is especially important for underrepresented populations, for which this type of analysis cannot be performed, and who are usually those who would benefit the most from research that could unravel, induced or endemic, inequalities of the population.

Lastly, although in the absence of information the parameters that we have estimated can be used to carry out several analyses in other parts of the world, including parameterizing epidemiological models, we have also seen that there can be significant differences across regions of a relatively small country. Thus, it is important to bear this in mind when comparing the effect of the pandemic in very different regions of the world, with diverse cultures and varying resource accessibility. The explosion on the availability of almost real-time data all over the world, represents a huge opportunity for the advancement of science and the education of society. Yet, at the same time, it shows that many of our information systems are still immature and that there is much work left to be done before we are ready for future pandemics.

## Data Availability

All the data is available in public repositories

https://cnecovid.isciii.es/covid19/

## Contributors

A.A, designed research with contributions from J.L.B, G.T.A and Y.M.; A.A performed research; A.A. and Y.M. analyzed the data and results; A.A., J.L.B, G.T.A. and Y.M. discussed results; and A.A. and Y.M. prepared the manuscript. All authors read and approved the final version. The funding sources had no role in the design, analysis, or writing of the manuscript.

## Data sharing

All the data and code necessary to reproduce the results will be available in https://github.com/aaleta/COVID_hospitalization and assigned a DOI upon publication. Due to confidentiality, the detailed data on the characteristics of hospitalized patients in Aragon cannot be shared, but all the analysis and the model estimation was carried out using publicly available data which can be accessed from the original sources, or in the GitHub folder.

