## Supplementary Material for "Unraveling the COVID-19 hospitalization dynamics in Spain using publicly available data"

**Estimation of the model parameters**

The prior and posterior distributions of the parameters for the region of Aragon are shown in figure S1. In table S1, we report the specific values obtained in the fitting, and figure S2 the corresponding distributions for the admission interval and the length of stay (LoS).

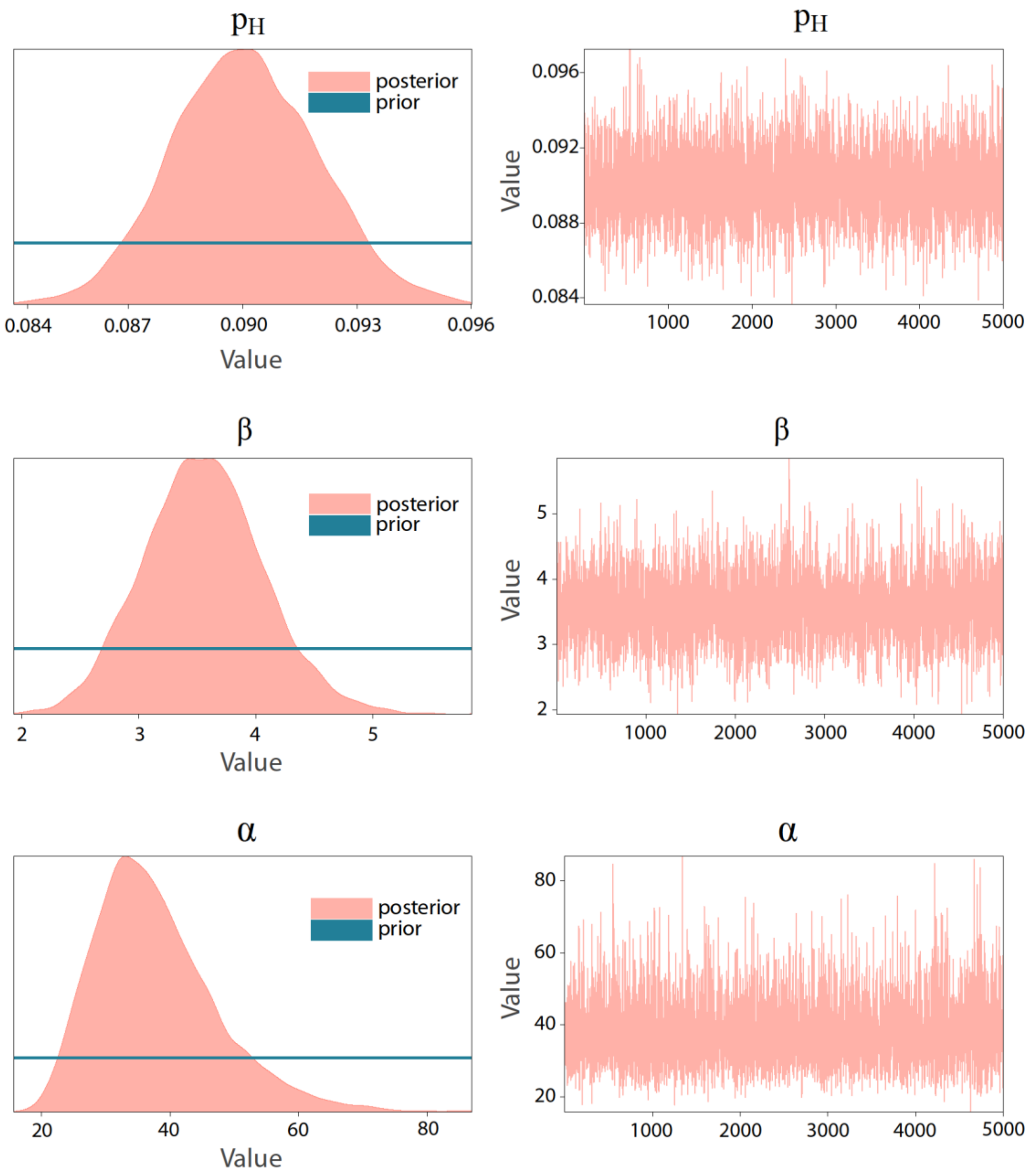

Figure S1: Prior and posterior distributions. Left column: prior and posterior distributions of the model parameters for new admissions in the region of Aragon. Right column: MCMC iterations. In all cases, the parameters were estimated for the region of Aragon using data from July 1 to December 1 2020 and their associated $\hat{R}$ was 1.0.

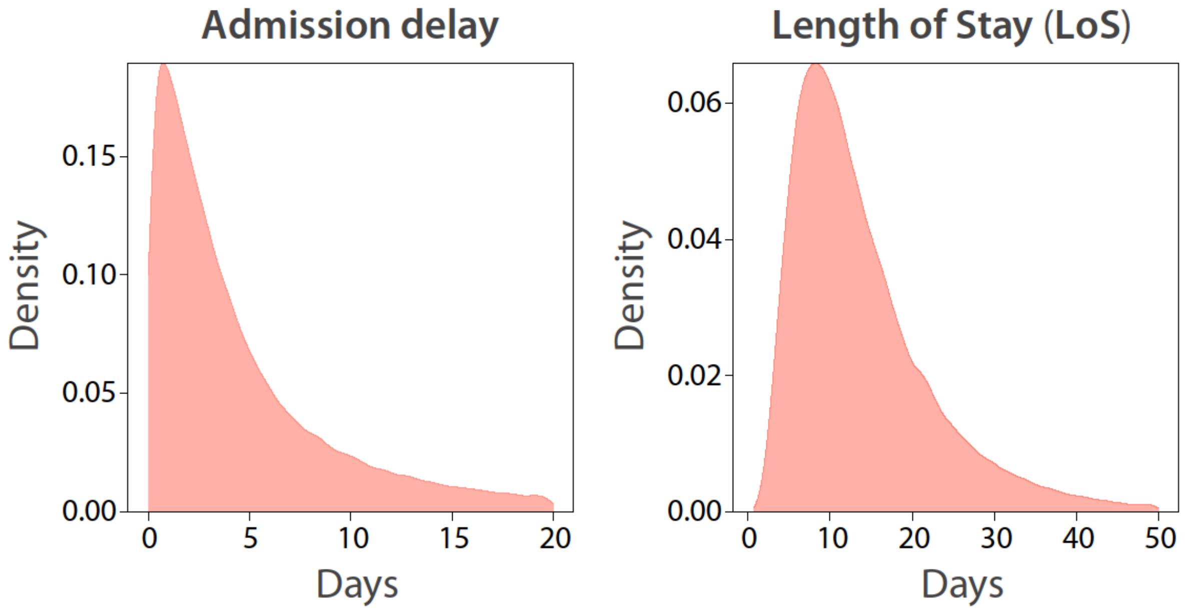

Figure S2: Time-interval distributions. Left: delay between symptom onset and hospital admission. Right: distribution of the Length of Stay (LoS) in the hospital.

| **Parameter** | **Mean** | **HDI 2·5%** | **HDI 97·5%** | $\hat{R}$ |
|---|---|---|---|---|
| $p_H^{AR}$ | 0·090 | 0·086 | 0·094 | 1·0 |
| $\beta^{AR}$ | 3·557 | 2·579 | 4·564 | 1·0 |
| $\mu^{AR}$ | 2·476 | 2·441 | 2·512 | 1·0 |
| $\sigma^{AR}$ | 0·620 | 0·554 | 0·688 | 1·0 |

Table S1: Estimated parameters for the region of Aragon. The probability of being admitted into the hospital upon being detected is $p_H$. The distribution governing the delay between case detection and admission is a Half-Cauchy distribution parameterized with $\beta$. The distribution on the length of stay (LoS) is modeled using a Log-Normal distribution parameterized with an average of $\mu$ and standard deviation $\sigma$.

**Surveillance data in Aragon**

In figure S3, we show the evolution of the reproduction number from July 1 2020 to December 1 2020 obtained from the incidence data provided by the regional government. In figure S4, we depict the age profile of the individuals admitted each week into any hospital in Aragon since vaccination started.

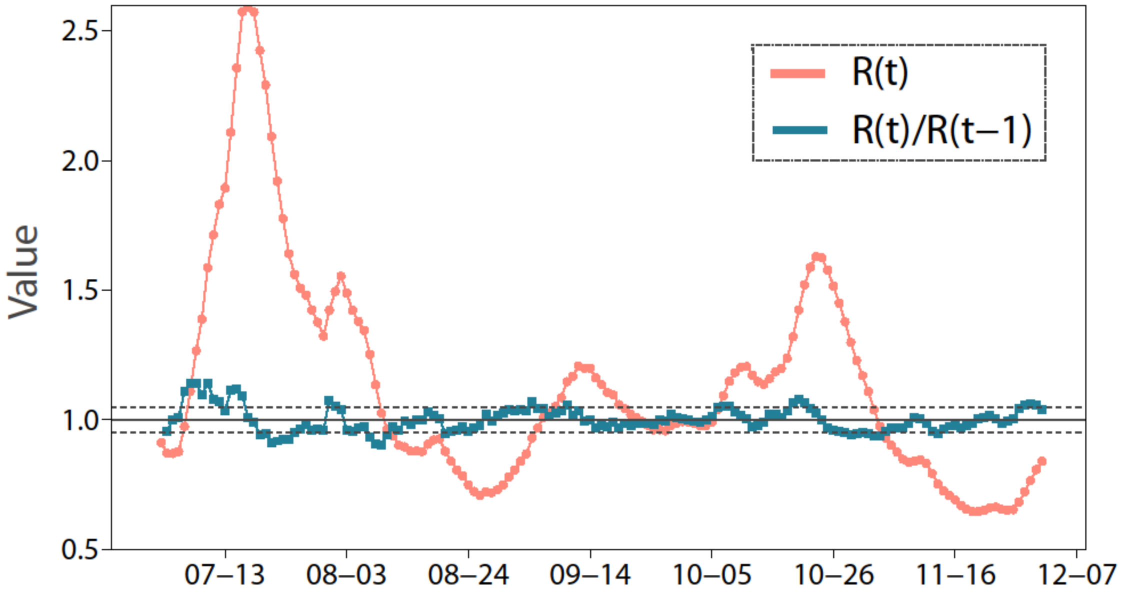

Figure S3: Estimated reproduction number as a function of time, $R(t)$, in Aragon. Circles represent the value of the reproduction number while squares show the quotient between the value at time $t$ over the value on the previous day, $R(t)/R(t-1)$. Dashed, horizontal lines represent the values of 0·95 and 1·05.

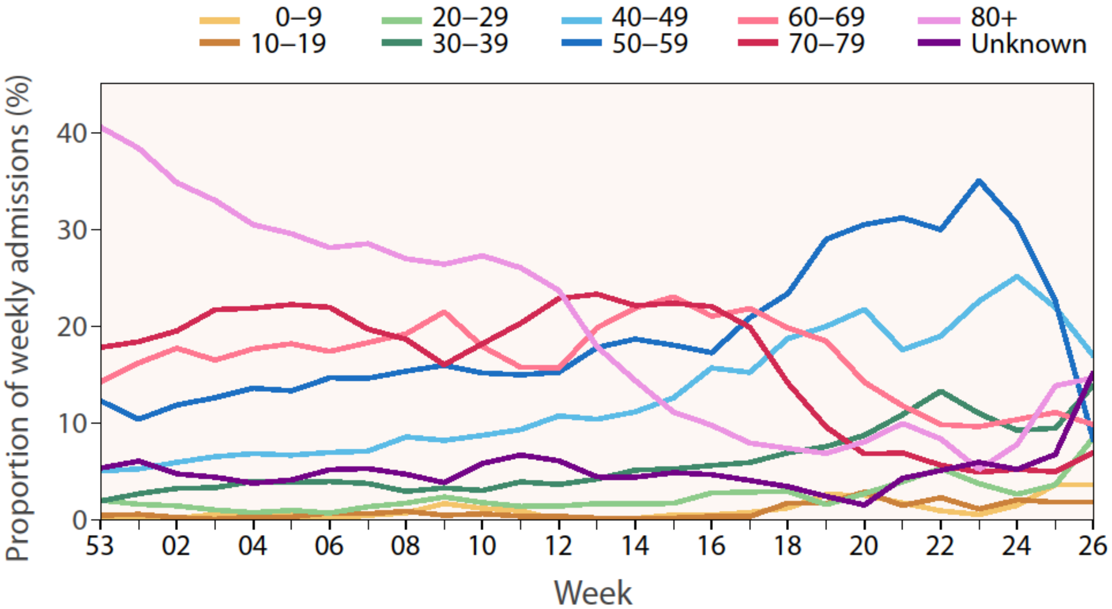

Figure S4: Age profile of hospital admissions. Proportion of weekly admissions corresponding to each age group in Aragon since vaccination started (week 53 of 2020). The curves have been smoothed using a rolling average of size 3.

**Regional results**

In figure S5, we show the number of new hospital admissions in each autonomous region in Spain, together with the prediction of the model. The MAPE value of the prediction for the third wave is presented in table S2.

| Autonomous Region | ISO Code | MAPE (%) | | | |
|---|---|---|---|---|---|
| | | January | February | March | April |
| Andalusia | AN | 6.94 | 8.87 | 7.58 | 6.65 |
| Aragon | AR | 5.60 | 5.27 | 5.09 | 4.71 |
| Principality of Asturias | AS | 59.54 | 61.59 | 53.47 | 49.69 |
| Canary Islands | CN | 7.41 | 7.00 | 7.51 | 7.51 |
| Cantabria | CB | 18.98 | 18.18 | 19.43 | 20.47 |
| Castille-La Mancha | CM | 10.48 | 7.27 | 5.63 | 4.37 |
| Castille and León | CL | 5.01 | 4.53 | 3.93 | 4.09 |
| Catalonia | CT | 4.79 | 5.36 | 4.61 | 4.08 |
| Extremadura | EX | 36.65 | 35.93 | 31.29 | 29.17 |
| Galicia | GA | 2.47 | 2.63 | 2.37 | 2.37 |
| Balearic Islands | IB | 15.43 | 10.15 | 10.59 | 12.60 |
| La Rioja | RI | 19.50 | 17.46 | 14.77 | 13.50 |
| Community of Madrid | MD | 20.13 | 30.38 | 32.27 | 32.02 |
| Region of Murcia | MC | 13.84 | 19.62 | 17.13 | 15.44 |
| Chartered Community of Navarre | NC | 9.92 | 7.94 | 6.96 | 6.22 |
| Basque Country | PV | 7.30 | 6.34 | 7.40 | 8.12 |
| Valencian Community | VC | 8.97 | 12.72 | 13.88 | 15.20 |

Table S2: Forecast quality in each autonomous region. For each region, the MAPE is estimated using the cumulative number of admissions up to the month represented in the table. In regions with high MAPE might have undergone changes in their contact tracing and testing procedures either during the training process or during the prediction phase.

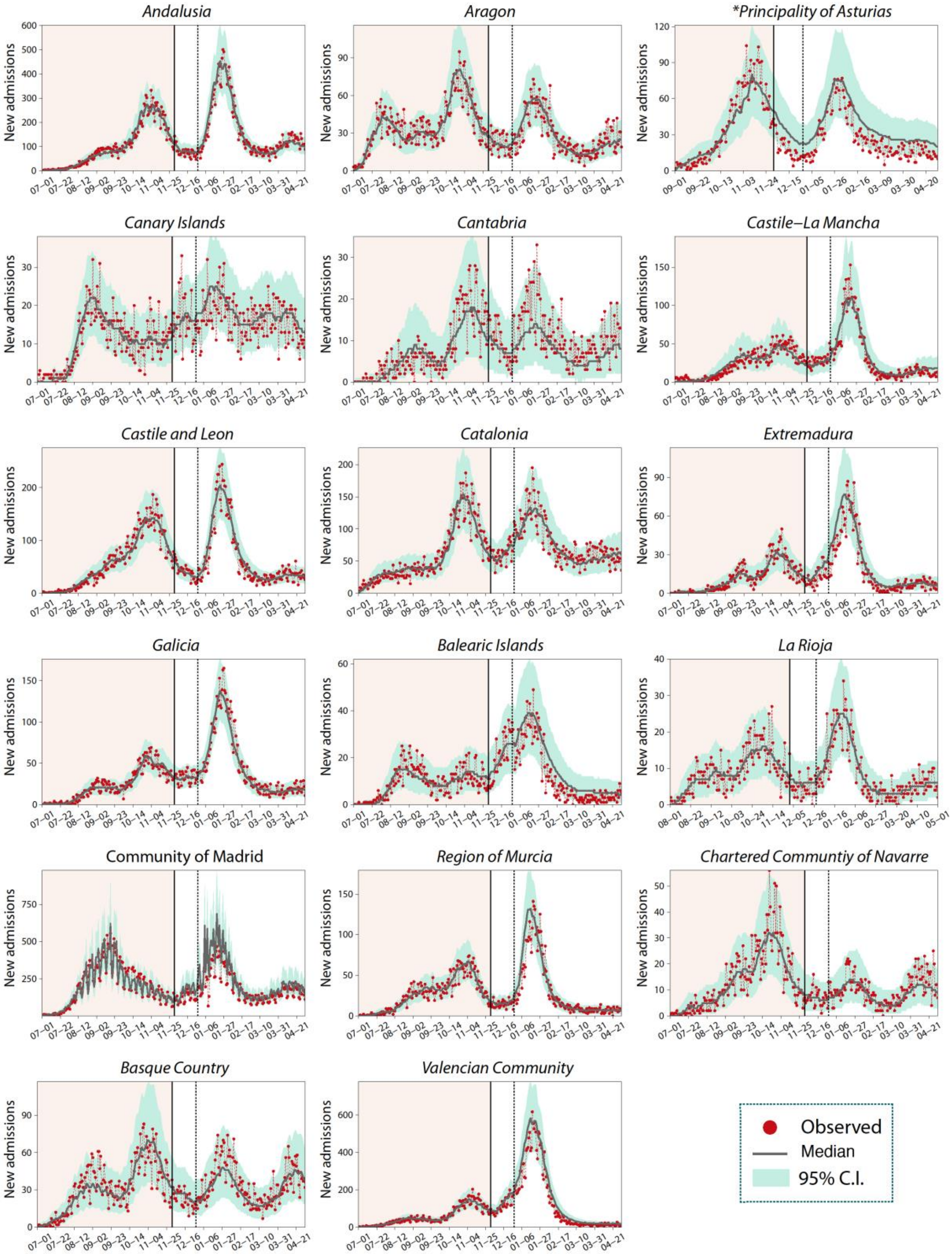

Figure S5: Number of daily admissions to hospital in each autonomous region. To estimate the parameters of the model, we use the information before December 1 2020. From December 1 the number of admissions is obtained using the estimated parameters for each region and the daily incidence. The solid line indicates the end of the training period and the dashed line the beginning of vaccine roll-out.

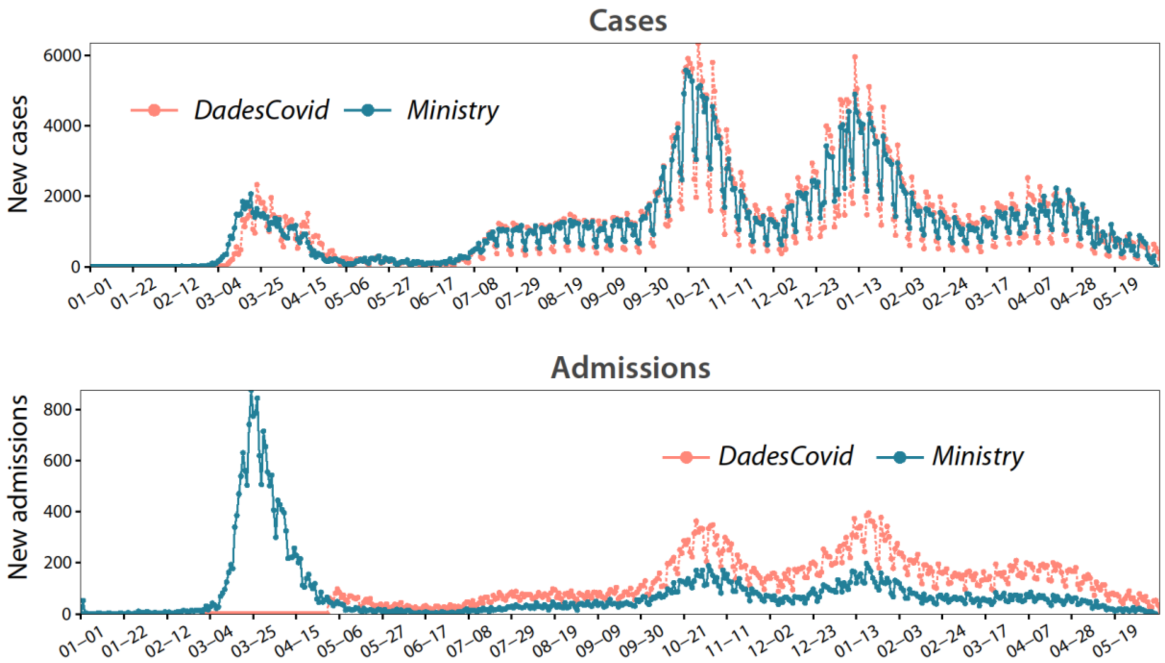

Figure S6: Comparison between the regional and national data for Catalonia. Top: number of new cases reported by DadesCovid (open data portal of the regional government) and by the Spanish Ministry of health. Bottom: number of new hospital admissions. Even though the number of new cases reported by both institutions agree, the number of new hospitalizations reported by the regional government is twice as large as the one reported by the ministry.

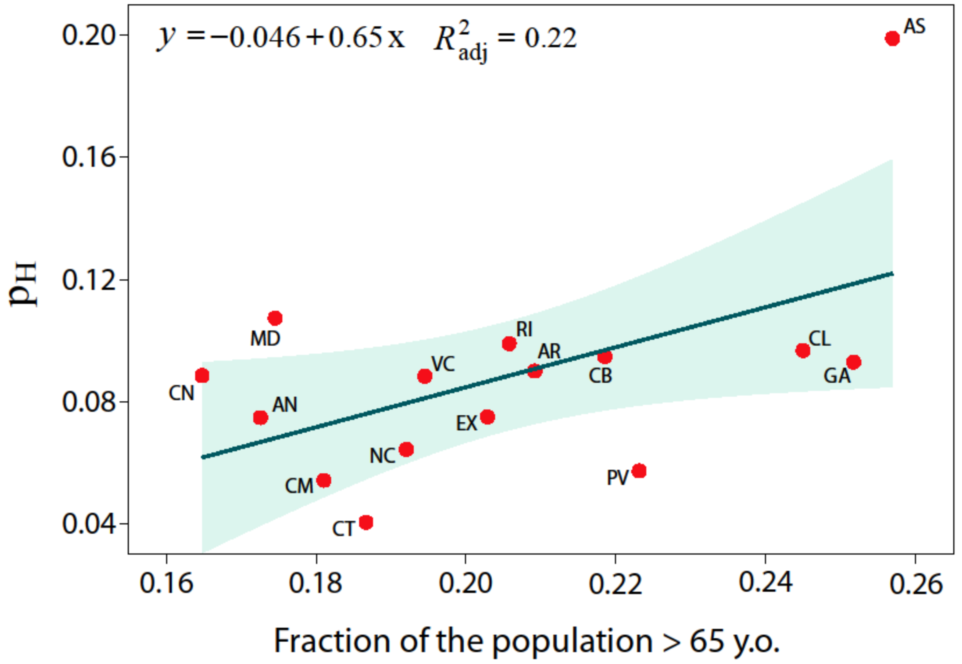

Figure S7: Effect of the regional age distribution on the admission probability. Correlation between the estimated value of $p_H$ in each region and the fraction of the population older than 65 years old. We obtain $R^2 = 0{\cdot}22$ signaling that the regression is not able to explain these differences. Furthermore, if we remove the data from the Principality of Asturias (AS) the correlation disappears completely.
